# Cancer Serum Atlas supported precise pan-targeted proteomics enable multi-cancer detection

**DOI:** 10.1101/2022.08.09.22278527

**Authors:** Anqi Hu, Lei Zhang, Zhenxin Wang, Chunyan Yuan, Ling Lin, Jiayi Zhang, Xia Gao, Xuguang Chen, Wei Guo, Pengyuan Yang, Huali Shen

## Abstract

The wide dynamic range of serum proteome restrained discovery of the clinically interested proteins in large cohort studies. Herein, we presented a high-sensitivity, high-throughput and precise pan-targeted serum proteomic strategy for high-efficient cancer serum proteomic research and biomarker discovery. We constructed a resource of over 2000 cancer-secreted proteins and the standard MS assays and spectra of at least one synthetic unique peptide per protein were acquired and documented (Cancer Serum Atlas,www.cancerserumatlas.com). Then, the standard peptides anchored parallel reaction monitoring (SPA-PRM) method was developed with support of Cancer Serum Atlas, achieving precise quantification of cancer-secreted proteins with high throughput and sensitivity. We directly quantified 325 cancer-related serum proteins in 288 serum of four cancer types (liver, stomach, lung, breast) and controls with the pan-targeted strategy, and discovered considerable potential biomarkers benefit for early detection of cancer. Finally, a proteomics based multi-cancer detection model was built, demonstrating high sensitivity (87.2%), specificity (100%), with 73.8% localization accuracy for an independent test set. In conclusion, the Cancer Serum Atlas provides a wide range of potential biomarkers that serve as targets and standard assays for systematic and high-efficient serological studies of cancer, and the Cancer Serum Atlas supported pan-targeted proteomic strategy enables high-efficient biomarker discovery and multi-cancer detection, thus can be a powerful tool for liquid biopsy.

## Introduction

Large-scale serum/plasma proteomics in population studies provides opportunities to capture markers of lifestyle and environmental exposure, to stratify individuals according to their state of health (or disease), and to monitor the longitudinal progression of disease(Geyer et al., 2017; Suhre et al., 2021). The challenge lies in quantifying proteins across a wide dynamic range, especially for large-scale samples(Suhre et al., 2021). The widely used data-dependent acquisition (DDA) MS methods or the emerging data-independent acquisition (DIA) approaches allow measurement of 300-450 proteins in hundreds of plasma samples(Geyer et al., 2016a; Geyer et al., 2016b; Liu et al., 2015; Messner et al., 2020),however, most of the identified proteins are high-abundant functional plasma proteins, which is difficult to achieve required specificity for targeted diseases.

Target mass spectrometry can reduce the complexity of the serum proteome caused by high abundant proteins and owns the advantages of high sensitivity and accuracy(Marx, 2013; Picotti and Aebersold, 2012). Recent progress of the emerging mode termed prmPASEF (parallel reaction monitoring-Parallel Accumulation-Serial Fragmentation) further achieved high resolution, sensitivity, and acquisition speed(Lesur et al., 2021). However, it is only applicable to specific target proteins and the throughput are relatively low, and thus its application is limited to biomarkers’ validation. Constructing a universal and clinically valuable target protein library while further improving the throughput and performance of the targeted proteomics would make this technology promising in large cohort studies to advance the protein biomarkers’ discovery and disease diagnosis.

The secretome is considered a valuable reservoir of potential biomarkers for cancer and other diseases(Basisty et al., 2020; Huettenhain et al., 2019; Robinson et al., 2019; Xie et al., 2020). In the past few years, clinical proteomic/proteogenomic research has produced a large number of high-quality cancer proteomics datasets(Chen et al., 2020; Clark et al., 2019; Dou et al., 2020; Gao et al., 2019; Ge et al., 2018; Gillette et al., 2020; Jiang et al., 2019; Li et al., 2020; Mun et al., 2019; Pozniak et al., 2016; Vasaikar et al., 2019; Xu et al., 2020). Filtering cancer-secreted proteins which release into a common distant medium, like blood, from differentially expressed proteins (DEP) of the published datasets, were expected to generate a valuable candidate biomarker library for various cancer research.

Multi-cancer detection offers the opportunity to screen multiple cancer types in population, thus identifying tumors when cures are more achievable(Ahlquist, 2018). Yet, such an approach would require high specificity, sensitivity, and accurate tissue of origin (TOO) identification(Ahlquist, 2018). Current multi-cancer detection studies are almost blood tests focusing on DNA. The Circulating Cell-free Genome Atlas Consortium used methylation signatures in cell-free DNA (cfDNA) to detect more than 50 cancer types across stages (Galleri)(Klein et al., 2021; Liu et al., 2020). CancerSEEK is another blood test for multi-cancer detection that can detect eight common cancer types through assessment of the levels of known protein biomarkers and mutations in cfDNA(Cohen et al., 2018).To date, there is no proteomic-based strategy for multi-cancer detection.

In this study, we developed a high-sensitivity, high-throughput and precise pan-targeted serum proteomic strategy for efficient cancer serum proteomic research and biomarker discovery. The strategy is supported with the Cancer Serum Atlas database we constructed (www.cancerserumatlas.com) that containing over 2000 cancer-secreted proteins for targets, as well as the standard peptide anchored PRM technology (SPA-PRM) enabling high-throughput and accurate PRM analysis. The Cancer Serum Atlas database contains cancer-secreted proteins filtered from published data of seven representative cancer types (liver, stomach, lung, breast, colorectum, kidney, and endometrium). What’s more, at least one unique peptides of each protein were synthesized and their retention time, MS parameters, and MS spectra were presented in the database as reference standard. Then, we developed the SPA-PRM assay. Through anchoring retention time (RT) window according to the RT of those standard peptides provided in Cancer Serum Atlas, the SPA-PRM enabled high-throughput and accurate pan-targeted proteomics that allow identifying and quantifying up to 2000 proteins in one run with sub-attomole sensitivity. The precise pan-targeted serum proteomic strategy can be directly applied to large cohort serum proteomic studies of various types of cancers without the discovery stage (differential proteome analysis). We then applied this strategy to directly quantify 400 cancer secreted proteins in sera of 288 individuals with four different types of cancer (liver, stomach, lung, breast) and normal controls. Without the discovery stage of the routine route, the Cancer Serum Atlas supported precise pan-targeted proteomics strategy accelerates the progress of biomarker discovery and clinical transformation. We found potential biomarkers that benefits (early) detection of cancer and the proteomics data were successfully applied to build a multi-cancer detection model, achieving high sensitivity (87.2%) and specificity (100%), with 73.8% localization accuracy for an independent test cohort. To sum up, the Cancer Serum Atlas supported pan-targeted proteomics strategy provided a powerful tool for serological studies of cancer, and the proteomics-based multi-cancer detection model demonstrated the high sensitivity, specificity, and localization accuracy with extremely low sample consumption, which is clinically attractive.

## Results

### Study design

In order to establish a pan-targeted serum proteomic strategy for cancer research, we firstly constructed Cancer Serum Atlas database that serve as the cancer-secreted proteins bank for target protein/peptide screening and the reference MS assay library for the SPA-PRM method development. In detail, we mined the differentially expressed proteins (DEPs) between tumor and non-tumor adjacent tissues of seven representative cancer types from the latest proteomic/proteogenomic research on large cohort patients (Chen et al., 2020; Clark et al., 2019; Dou et al., 2020; Gao et al., 2019; Ge et al., 2018; Gillette et al., 2020; Jiang et al., 2019; Li et al., 2020; Mun et al., 2019; Pozniak et al., 2016; Vasaikar et al., 2019; Xu et al., 2020) **(Figure 1)** and further filtered 2169 potentially secreted DEPs according to the results of GO categories, signalP prediction and preliminary screen in pooled serum. Then, 2700 unique peptides of the potentially secreted DEPs were synthesized and the optimized LC-MS parameters and their high-quality MS/MS, MRM, and 4D-PRM spectra were acquired and collected as standard reference. After screening the detectable proteins from the Cancer Serum Atlas as target protein list, SPA-PRM method was developed on the basis of the high-sensitivity prmPASEF technology. The analyte throughput was increased by the scheduled PRM that anchoring the RT window to ±1min of the standard peptide’s RT recorded in the database, and accuracy of the peptide identification was ensured through referencing standard MS spectra in the Cancer Serum Atlas. We then applied this strategy to directly quantify 400 cancer-secreted proteins in sera of 288 individuals of 74 controls (C), 52 hepatocellular carcinomas (HCC), 57 gastric cancer (GC), 56 lung cancer (LC), and 49 breast cancer (BC), thus constructed a multi-cancer detection model **(Figure 1)**.

**Figure 1.**
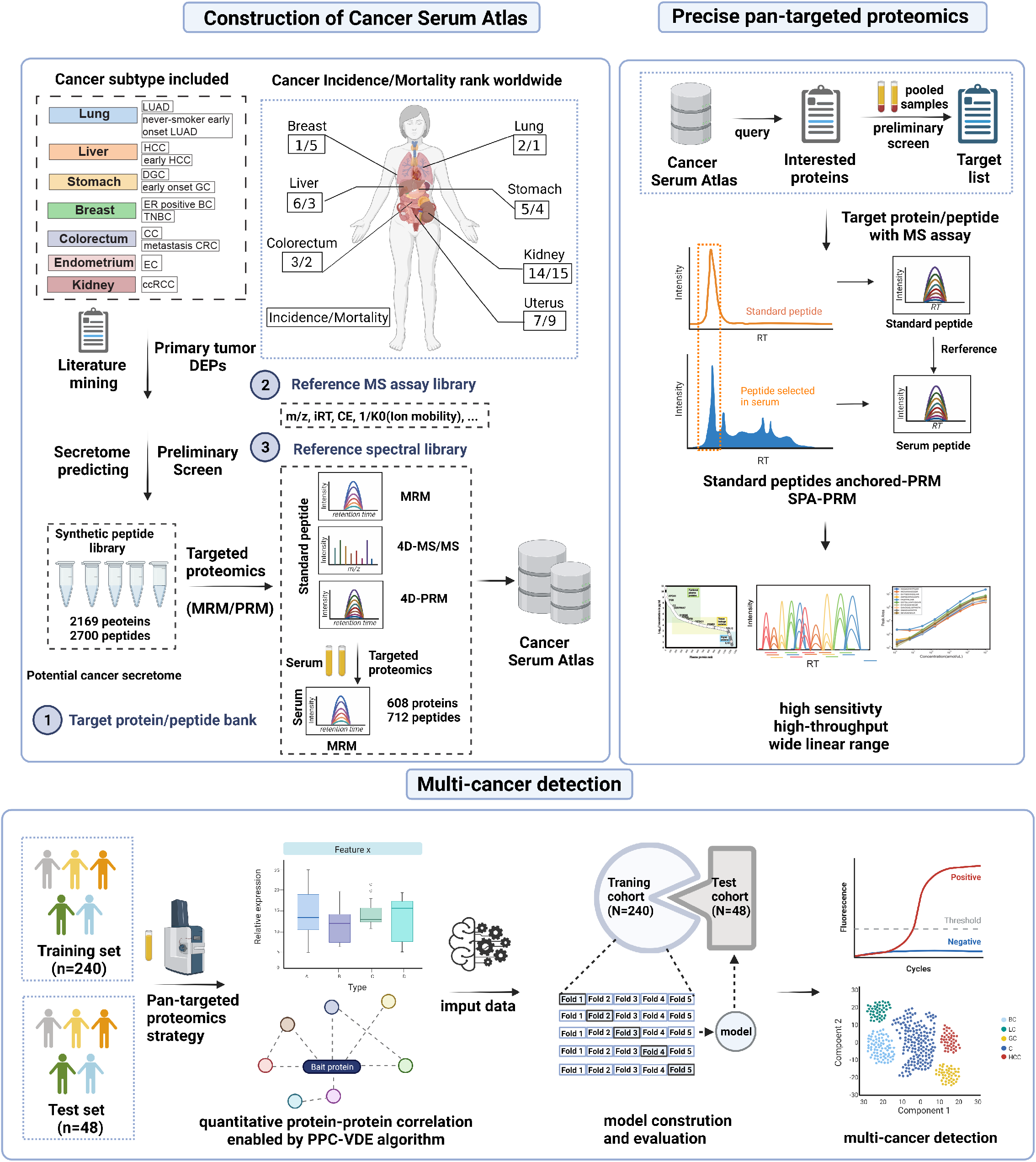
Schematic illustration of study design. LUAD, Lung adenocarcinoma; DGC, diffuse-type gastric cancer; ER, estrogen receptor; TNBC, triple-negative breast cancer; CRC, colorectal cancer; EC, endometrial carcinoma; ccRCC, clear cell renal cell carcinoma; LC, lung cancer; GC, gastric cancer; HCC, hepatocellular carcinoma; BC, breast cancer; PPC-VDE, protein-protein correlations based variable dimension algorithm; DEP, differentially expressed protein; MRM, multiple reaction monitoring; PRM, parallel reaction monitoring. SPA-PRM, standard peptides anchored-parallel reaction monitoring.

### Pan-cancer analysis and generation of cancer secreted protein list

Proteins secreted by cancer cells release into a common distant medium, like blood, making it a valuable candidate library for serum biomarker discovery. We constructed cancer-secreted protein database in Cancer Serum Atlas by filtering DEPs through differential proteome analysis of primary tumor and paired-normal tissue from the latest large-scale human tumor proteomics researches. The first version of Cancer Serum Atlas including data of 7 cancer types (liver, stomach, lung, breast, colorectum, kidney, and endometrium) are derived from large cohort cancer proteomic studies published before Jun., 2020[14-23, 25] with the proteomic data of paired tumor and non-tumor adjacent tissues. However, the data of colorectal cancer was incomplete (only 35 DEP data)[23]. The database was continuously updated and has been updated once to include high-quality proteomic data before Nov., 2020 [24]. The current version of Cancer Serum Atlas includes data from seven representative cancer types (liver, stomach, lung, breast, colorectum, kidney, and uterus) consisting of twelve subtypes **(Figure 1)**, These seven cancer types had high incidence and mortality, which taking together accounts for more than 50% of all cancer types worldwide (http://gco.iarc.fr/) **(Figure S1A)**. We filtered 9095 DEPs according to the literature **(Figure 2A-B, Table S1-S2, Methods)**, in which more than 50% DEPs showed cancer type-specificity. GO-biological process analysis revealed that the dysregulated proteins of each cancer type were closely related to the primary tissue’s function (**Figure S1B)**. Through bioinformatic analysis (Signal P and GO-CC, **Methods**), 4062 proteins were categorized or predicted as secreted proteins **(Table S2)**, accounting for about 57% of the human secretome(Uhlen et al., 2019). It is noteworthy that the Signal P and GO-CC strategies for secreted proteins prediction are complementary to each other **(Figure S1E)**. Compared to the previous study, we adopted relatively less stringent filtering criteria to involve as many potential cancer-secreted proteins as possible. **Figure S1C**-**S1D** shows the detailed information for GO-CC analysis. Furthermore, the growth trend of cancer-secreted proteins significantly slowed down with increasing cancer types, suggesting that most of the cancer-secreted proteins may have been included in the database **(Figure S2A)**. Although these data were derived from seven cancer types, they achieved deep coverage of cancer-secreted proteins. **Figure S2B** shows the overlap of secreted proteins of 7 cancer types.

**Figure 2.**
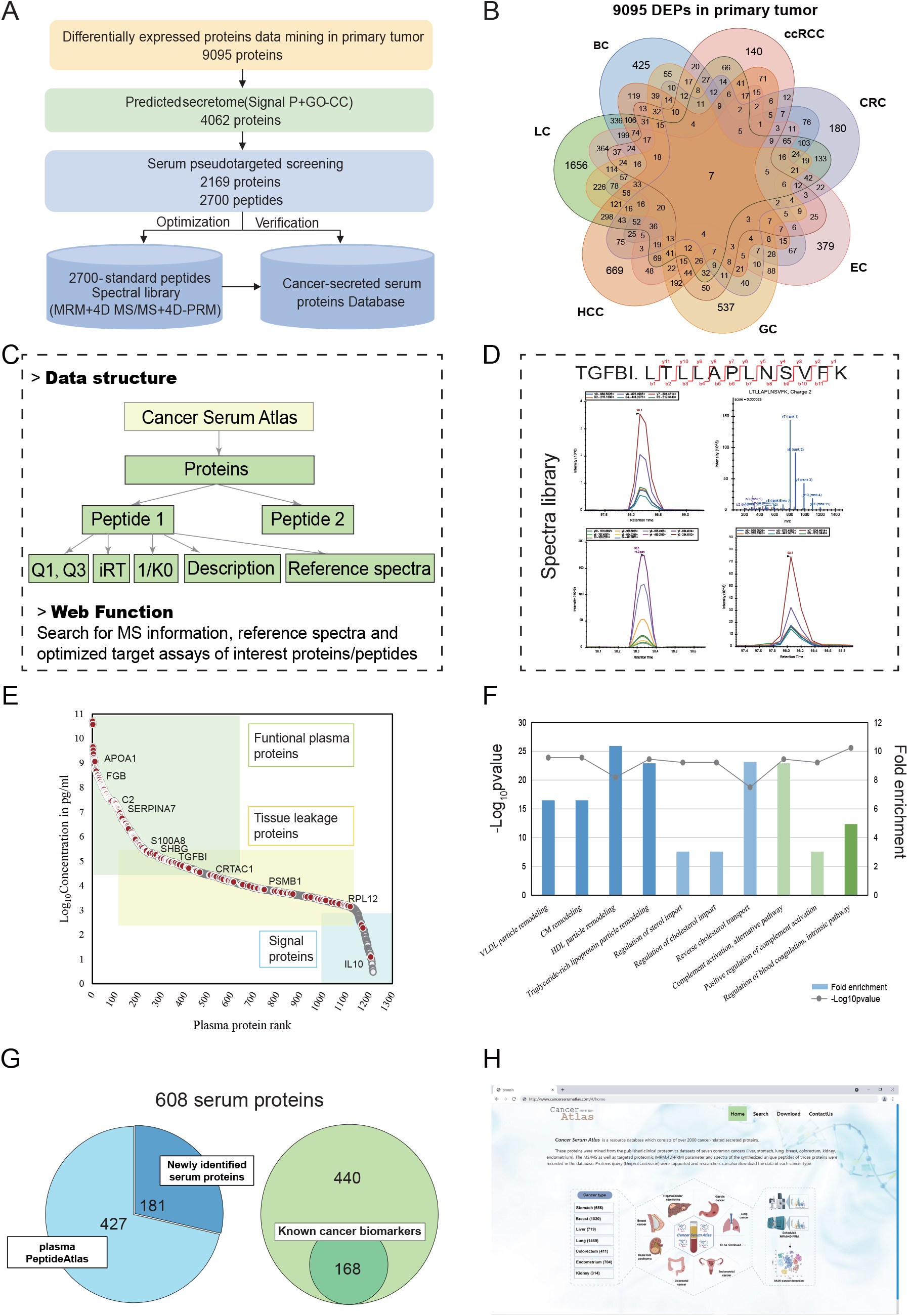
Overview of Cancer Serum Atlas. (A) A schematic diagram of Cancer Serum Atlas data collection. GO-CC, gene ontology-cellular component. Signal P, a signal peptide prediction software. (B) Distribution of the 9095 differentially expressed proteins (DEPs) of seven cancer types. (C) The data structure and function of Cancer Serum Atlas (website). (D) A typical example of MS spectra exhibited in the website of peptide (LTLLAPLNSVFK) for the TGFBI protein. (E) Abundance distribution of 608 serum proteins (red dots) according to the reference concentrations in the human plasma proteome database (http://www.plasmaproteomedatabase.org/). (F) The serum proteins-related biological processes analyzed by Gene-Ontology. (G) The 2D pie chart shows the composition of the serum proteins. (H) Display of the Cancer Serum Atlas website.

### Construction of the Cancer Serum Atlas

The 4062 putatively cancer-secreted proteins **(Table S2)** were preliminarily screened in pooled serum using the pseudo-targeted LC-MRM method(Xie et al., 2020). As a result, the 2700 detected peptides of 2169 proteins were filtered for further Cancer Serum Atlas construction **(Figure 2A)**.The reference MS assays of unique peptides (standard peptides) and detectable serum peptides of target proteins were also included in Cancer Serum Atlas. The 2700 preliminary identified unique peptides were chemically synthesized, their LC-MRM or PRM parameters were optimized, and the MS/MS, MRM, and 4D-PRM spectra were acquired. Well-organized information including protein, peptide (sequence), reference spectra of the standard peptides (MRM, 4D-MS/MS, 4D-PRM), as well as the MRM/PRM parameters (Q1; Q3; 1/K0; iRT; CE; description) were integrated to construct the library **(Figure 2C)**. iRT value provides standard convertible retention time. **Figure 2D** shows the content of the spectral library of the Cancer Serum Atlas, taking the peptide LTLLAPLNSVFK of TGFBI as an example. Proteins or peptides queries were supported. Researchers can also download the data of each cancer type for establishment of targeted proteomic assays or checking the accuracy of peptide identification. Then, we detected the 2700 peptides of 2169 cancer-secreted proteins in eight pooled serum digests of seven cancer types (liver, stomach, lung, breast, colorectum, kidney, and endometrium) and normal control respectively using the scheduled MRM method **(Table S3**) described in our previous study(Xie et al., 2020). A total of 712 unique peptides corresponding to 608 proteins obtained experimental evidence in serum samples. The dynamic range of the 608 serum proteins spanned more than 10 orders of magnitude according to their estimated concentration in the plasma proteome database published in May 2017 (http://www.plasmaproteomedatabase.org/) **(Figure 2E)**. These proteins mainly function in lipoprotein remodeling (blue), regulation of lipids metabolic process (light blue), complement activation (light green), and regulation of blood coagulation pathway (green) **(Figure 2F)**. Compared with the Plasma Atlas built by Human Proteome Organization’s Human Plasma Proteome Project (HPPP), we newly identified 181 serum proteins **(Figure 2G)**. In addition, more than 160 cancer biomarkers described in previous studies were included in our dataset **(Figure 2G)**. Cancer Serum Atlas demonstrated the deep coverage of the cancer-secreted proteins in serum. The spectra of the 712 peptides detected in serum were also recorded in Cancer Serum Atlas. The resource is available for download from the online website (www.cancerserumatlas.com) **(Figure 2H)**.

### SPA-PRM method for pan-targeted cancer serum proteomics

Recent progress of prmPASEF achieved high sensitivity, and acquisition speed(Lesur et al., 2021). However, implementation of this method requires the identification of a list of target proteins, so the discovery phase using DDA/DIA methods is inescapable. In addition, the throughput is generally tens to one hundred peptides per run. The Cancer Serum Atlas can provide target protein list for the pan-targeted proteomics strategy applying to various cancer serum study. Furthermore, the database provides the standard RT and other MS assays for all the cancer-secreted proteins’ unique peptides that help anchoring target peptides in serum samples and establish scheduled PRM methods to significantly increase analytical throughput. Taken together, with the Cancer Serum Atlas’ supporting and on the basis of the SPA-PRM method, we proposed the precise pan-targeted proteomics strategy for cancer research in serum. This strategy replaces the regular discovery phase using DDA or DIA methods with a simple one-step screening experiment, and enables precise and accurate PRM quantification/verification of hundreds of proteins in large cohort study. Thus, the Cancer Serum Atlas supported precise pan-targeted proteomics strategy can expand the application to simultaneous biomarker discovery and validation in large cohort study.

We first optimized the prmPASEF assay with standard peptides. Through optimizing accumulation time durations, retention time window and LC gradient, the prmPASEF assay demonstrated an LLOD of 0.25 amole **(Figure 3A)** with more than 3-fold signal-to-noise ratios **(Figure 3B)** and an LLOQ of 1.28 amole **(Figure 3C)**. The linear range of most standard peptides spaces 4-5 orders of magnitude **(Figure 3D)**. After optimization, the sensitivity and linear range remarkably improved than the manufacturer’s(Lesur et al., 2021). In the analysis of 240ng serum digest, the fine-tuned system achieved narrow peak widths **(Figure 3F)** and stable retention time (pre-set time ± 0.2min) **(Figure 3G)** with numerous samples continuous injection. With such a stable system, all the target peptides can be successfully captured in a detection window of 2 min (preset RT±1 min) and the vast majority of peptides can obtain 10 to 20 data points that ensure accurate quantification **(Figure 3H)**. Thus the maximum throughput of a scheduled PRM assay with an detection window of each peptide as 2 min could achieve 2700 proteins per run **(Figure 3I)**, which is sufficient to match the current detection ability of serum proteomics. If the RT window is further narrowed, the detection throughput can be further improved **(Figure 3I)**.

**Figure 3.**
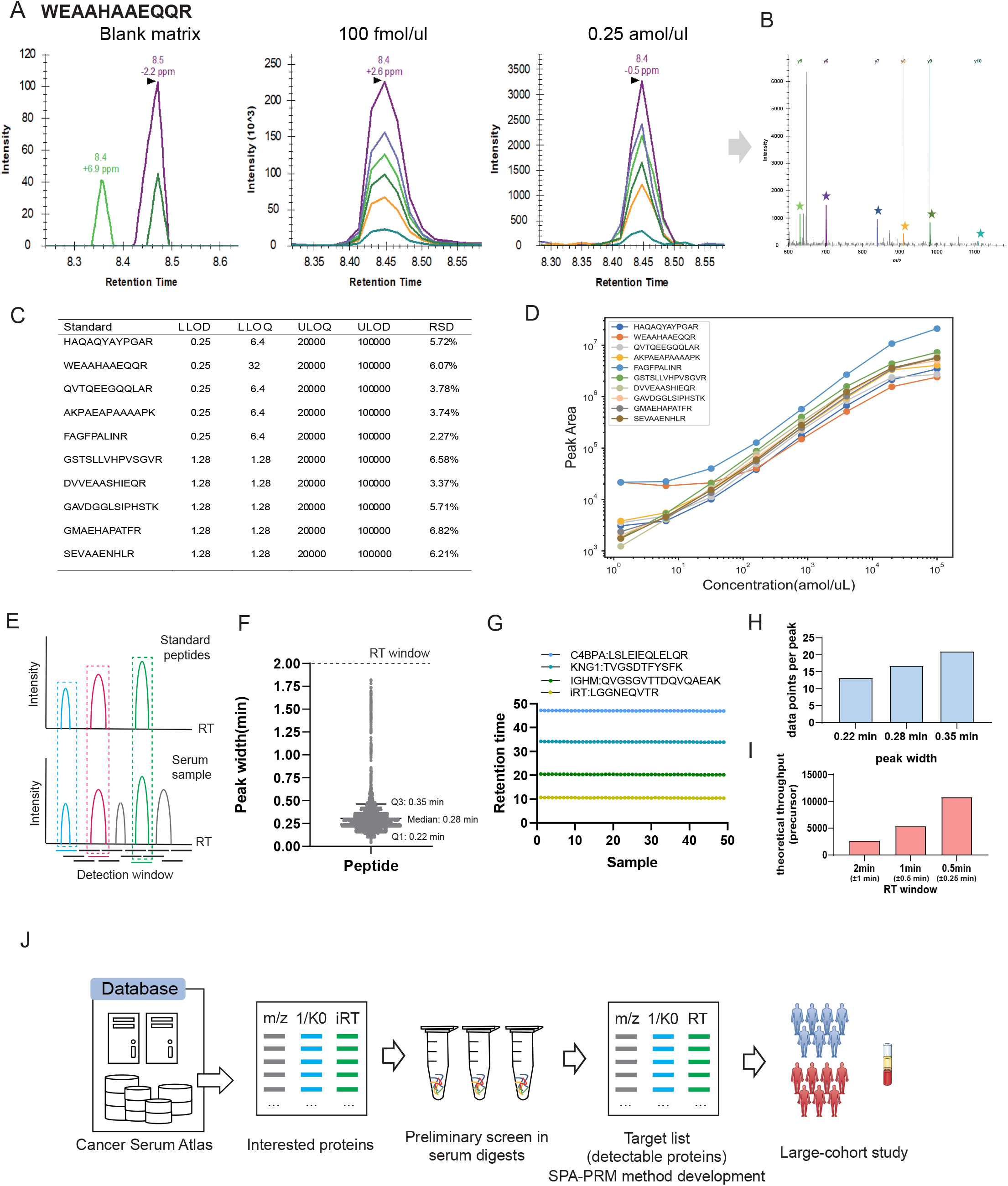
Performance of the SPA-PRM method. (A) Representative LC-prmPASEF spectra of the peptide WEAAHAAWQQR at different quantity. (B) MS2 spectrum of the peptide WEAAHAAWQQR at 0.25 amole. (C) Limits of detection and limits of quantification for standard peptides. (D) The linear range of the assay. (E) Schematic of SPA-PRM. (F) The distribution of the peak width of the detected serum peptides. (G) The stability of the retention time of 4 representative peptides across the 50 runs. (H) Practical data points per peak acquired in serum samples. (I) The theoretical throughput of the SPA-PRM when fixing TIMS accumulation time at 100ms according to different RT windows in our study. (J) Workflow of the pan-targeted proteomics strategy.

We then developed a pan-targeted proteomics strategy with the support of Cancer Serum Atlas **(Figure 3J)**. Firstly, filter the interested proteins from the Cancer Serum Atlas. Secondly, screen the interested proteins in pooled serum digests to generate a target list of detectable proteins. Then, the SPA-PRM method was set up according to the RT, 1/K0, and m/z of the standard peptides for large-cohort study. Through anchoring serum peptides to the standard peptides **(Figure 3E)**, the SPA-PRM method achieved the high detection throughput and precise peptide/protein identification and quantification. To sum up, the Cancer Serum Atlas supported SPA-PRM method featuring high sensitivity, high throughput, and wide linear range, enabling directly accurate identification and quantification of numerous cancer-secreted proteins for large cohort studies.

### Direct quantification of cancer-related serum proteins in a large cohort with the pan-targeted proteomics strategy

We chose four representative cancer types (liver, stomach, lung, breast) from the first version of Cancer Serum Atlas for the precise pan-targeted proteomic analysis. The 2 types of cancer (kidney and endometrium) were excluded due to the relatively low incidence and mortality. They did not belong to the 10 most common cancer types with high incidence and mortality (https://gco.iarc.fr/today/home). The colorectal cancer was excluded due to the incomplete proteomic data (only 35 DEP data)(Li et al., 2020) at that time point. We collected a cohort of 288 serum samples containing 74 C, 52 HCC, 57 GC, 56 LC, and 49 BC. The representativeness of these 4 types of cancer is very strong, which has higher incidence and mortality worldwide. The clinical characteristics including age, sex, cancer type, TNM stage, TNM grouping, histopathology and values of classical tumor markers were summarized in **Figure S3A-S3B** and **Table S4**. Based on the clinical data **(Table S4)**, we calculated the positive rate of 7 classical tumor markers (AFP, CEA, NSE, CYFRA21-1/cytokeratin 19 fragment, SCC/squamous cell carcinoma antigen, CA15-3/carbohydrate antigen 15-3, CA 72-4/carbohydrate antigen 72-4) in each cancer type **(Table S5)**. AFP is the most sensitive marker with a positive rate of 54.9% in HCC patients, and the positive rates of the remaining markers are less than 25% **(Table S5)**, which indicated that the existing cancer markers were insufficient to meet the requirements of cancer detection.

We randomized all the samples and used iRT peptides, hela digests, pooled serum digests as quality control for minimizing batch effects **(Figure 4A)**. The developed SPA-PRM method was applied to quantify 490 target peptides from 400 proteins in these samples. Firstly, we filtered the detectable cancer-secreted proteins in five pooled serum digests (C, HCC, GC, BC, LC) to generate a target list from the Cancer Serum Atlas. After careful selection, 490 unique peptides corresponding to 400 proteins were included in the target list for the subsequent quantitative analysis. Then, the SPA-PRM method was set up according to the RT, 1/K0, and m/z of the 490 standard peptides with 2 min detection window (preset RT±1 min). Details on quality control and reproducibility are detailed in Supplementary **Figure S3C-S3F**. We quantified 409 peptides corresponding to 325 proteins. Across different sample groups, on average 295 proteins were quantified and a high degree of protein identification overlap among groups was observed **(Figure 4B)**. The data (270 peptides, 194 proteins) had less than 50% missing values across any group in the training cohort were included for further analysis **(Table S6)**. When comparing each cancer type with control, nearly 70% of proteins demonstrated significant changes (P<0.05), among which 7 proteins were newly identified beyond the Plasma Atlas and 79 proteins (58.5% of DEPs) were undiscovered biomarkers, further declaring that directly targeting proteins in the Cancer Serum Atlas is an effective way to find cancer-related serum proteins **(Figure 4C). Figure S3G** showed the detailed differentially expressed proteins in each cancer group versus the control. More proteins tend to be up-regulated, and a few proteins were down-regulated when comparing cancer versus control. Among them, GC and HCC groups have the most DEPs versus control, followed by LC, and the BC has the least DEPs. The heatmap displayed significant differences in cancer versus control and among cancer types **(Figure 4F)**. These DEPs belong to three major pathways, namely lipoprotein and cholesterol metabolic process, complement activation, and platelet degranulation **(Figure 4F)**. OPLSDA plots showed that normal controls were well isolated from the rest cancer samples **(Figure 4D)**. For cancer types, GC and HCC were resolved from the other with few overlaps, but the BC and LC share a certain degree of similarity **(Figure 4E)**. Then, we preliminarily compared the performance of our proteomic data with classical tumor markers in cancer detection. We used the Luminex liquid suspension chip to detect the classical tumor markers commonly used in clinic(Cohen et al., 2018; Duffy, 2013; Reiter et al., 2015; Sung and Cho, 2008; Zhu et al., 2020), which includes alpha fetoprotein (AFP), carcinoembryonic antigen (CEA), neuron-specific enolase (NSE) and osteopontin (OPN) in 75 serum samples (15 samples of each group) randomly selected from the 288 serum cohort. The proteomic data of the corresponding samples were used for comparison. We found 15, 11, 6, 5 proteins of HCC, GC, LC and BC from our proteomic data that had better diagnostic performance than classical tumor markers **(Table S7-S8)** The above results demonstrated that the pan-targeted proteomics strategy can be directly applied to analyze a large cohort of samples with simple screening step, and showed high efficiency in discovering cancer-related proteins or even cancer biomarkers.

**Figure 4.**
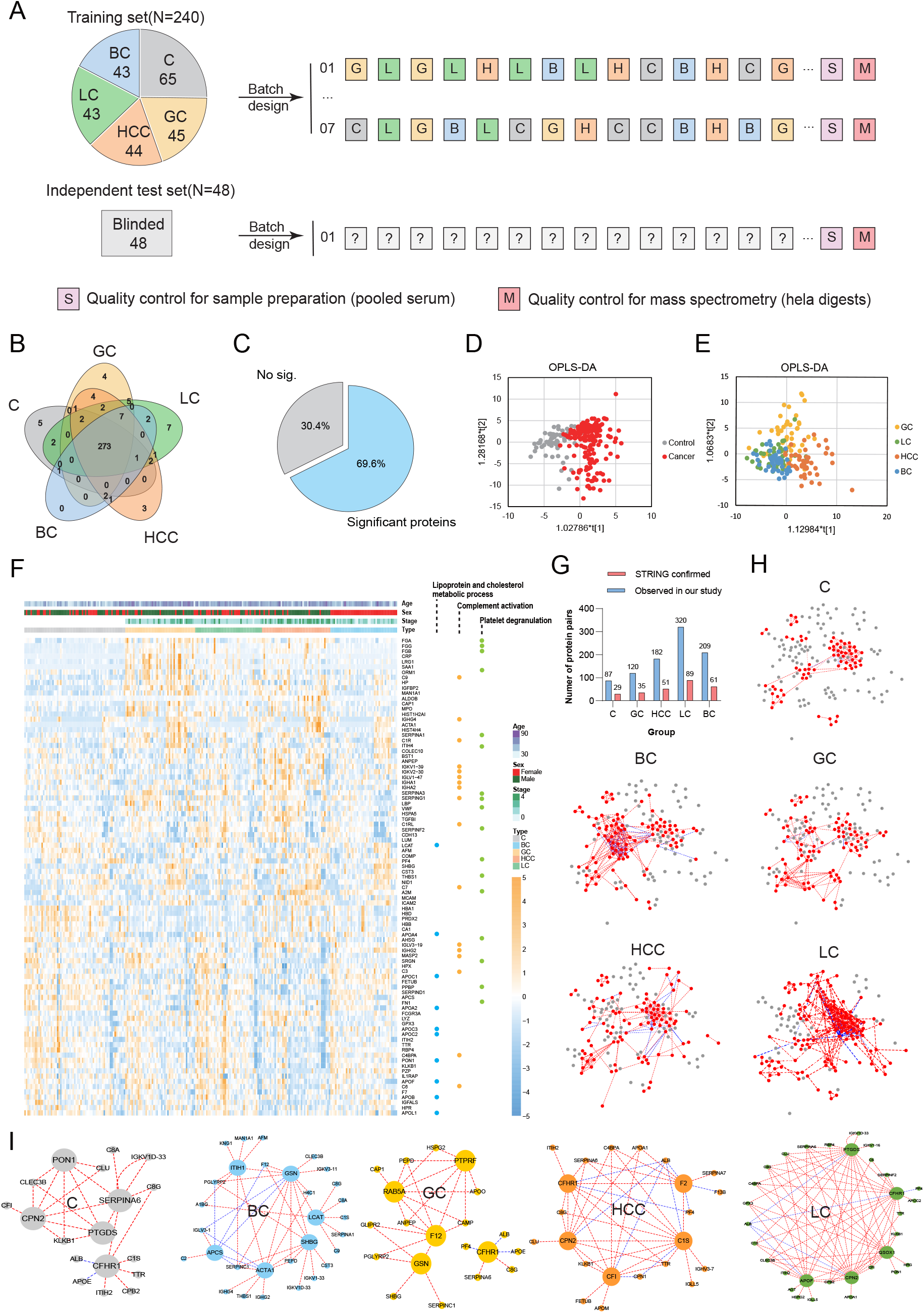
Cancer-related serum proteins quantification in a large cohort. (A) Batch design. (B) Venn diagram displaying the protein identification among five groups. (C) The 2D pie chart shows the percentage of significant proteins (cancer versus control, P-value<0.05). (D-E) The OPLS-DA models showing the separation of cancer and normal patients, as well as cancer types. (F) Heatmap of the DEPs within five groups (ANOVA, adjusted P-value<0.05) with the involved three pathways. (G) The number of correlated protein-protein pairs verified by STRING. (H) Protein-protein correlation networks. Proteins nodes in each group of samples are highlighted (red) attached with positive correlation (red edge) and negative correlation(blue) on the background of all the high protein-protein correlation networks for five groups (Pearson correlation coefficient more than 0.65 for each group were selected for total network construction). (I) The core co-regulatory network in each group (the nodes with the top 5 number of edges were selected).

### Protein-protein correlation analysis

Other than changes in protein expression, we also observed significant differences in protein-protein correlations (PPCs) across the five groups. The Pearson correlation coefficients between proteins were calculated to represent the protein-protein co-regulatory relationship. We found 632 highly correlated protein pairs (Pearson r>0.65) in the five groups totally, and about one-third has evidence in STRING **(Figure 4G)**. Then, we constructed an integrated network and highlighted the group-specific PPCs, respectively. PPCs across groups ranged from 87 in C (13.76%) to 320 in LC (50.63%) **(Figure 4H)**, indicating that not only the expression level of cancer-related proteins changed, but also the co-regulatory relationships were enhanced. We exhibited the core network of each group **(Figure 4I)**. It is noted that the core network (top 5 nodes with edges) of the five groups are different from each other. For instance, PON1 and SERPINA6 were specific in C, and QSOX1 and APOF were specific in LC. The core network of HCC was focused on a series of complement and coagulation factors. The differences of the core networks may instruct different characteristics of cancer types. Furthermore, some nodes (CFHR1, GSN, CPN2, etc) were shared within groups. Furthermore, we found 379 pan-cancer PPC pairs, the RAB5A-CAMP is a representative pan-cancer PPC pair. There were strong correlations from 0.69 to 0.92 between RAB5A and CAMP across all cancer samples **(Figure S4B)** whereas poor correlation (0.47) in normal controls **(Figure S4A)**. The effects of CAMP include activation of cell migration, proliferation, and invasion, regulation of cell apoptosis. Accumulating evidence indicates the significant role of CAMP in human cancer, with the tumorigenic or anti-cancer effects varied from cancer types(Chen et al., 2018). RAB5A also plays a vital role in tumorigenesis and distant metastasis, which positively regulated the extracellular signal-regulated kinase (ERK) signaling pathway and promoted epithelial-mesenchymal transition (EMT)(Yang et al., 2018; Zhang et al., 2017). So far, there is no evidence of the co-regulatory relationship of RAB5A-CAMP in the blood of cancer patients. Our results implied that the two proteins may have a co-regulatory relationship in cancer. All these results showed the fact that altered PPCs can infer vital differences within disease types. In addition, nearly half of PPCs in serum were observed in the primary tumor **(Figure S4C)**, and **Figure S4D-S4E** showed the different biological processes enriched in serum or tissue-specific networks, implying cancer-related serum proteomics has some unique characteristics compared with the primary tumor.

To further explore the cancer-specific characteristics and increase cancer-specific features, we applied the previously developed protein-protein correlation based variable dimension expansion (PPC-VDE) method(Xie et al., 2020) through implementing multiplication and division for every two peptides’ intensities and taking the resulted values as new variables **(Figure S4I)**. For example, HBB or MPO could not clearly separate the C and LC groups **(Figure S4F)**. While the HBB/MPO exhibited superior performance that significantly distinguished the two groups **(Figure S4G-H)**. Through dimension expansion, the number of significant features remarkably increased **(Figure S4J)**, especially for poorly distinguishable cancer types. The significant features (P<0.001) between LC and BC increased from 4 to 292 and the area under the ROC curve (AUC) was effectively improved from 0.765 to 0.886 **(Figure S4K-M)**. Beyond that, the AUCs of other hardly distinguishable groups (GC vs BC, C vs BC) were also noticeably increased after PPC-VDE **(Figure S4K-M)**, laying the foundation for subsequent feature selection and model construction.

### Biomarker discovery and multi-cancer detection enabled by the pan-targeted proteomic data

The pan-targeted proteomics strategy showed high efficiency in biomarker discovery. Nearly 70% of proteins demonstrated significant changes (P<0.05) when comparing each cancer type with control **(Figure 4C)**. We showed some representative serum biomarkers with potential diagnosis value **(Figure 5A-D)**. These proteins showed consistent trends of expression in tissues and serum when comparing cancer versus controls. For instance, the IGFALS protein was significantly down-regulated in HCC serum samples **(Figure 5A)**, which was also confirmed at the tissue level. Qiang Gao et al. had detected the down -regulation of IGFALS in tumors comparing with non-cancerous adjacent tissues (NATs) of HCC(Gao et al., 2019). The same for MCAM, IGFBP2, and LRG1 protein, as potential serum biomarkers identified in our study **(Figure 5B-D)**, they were also detected in tumor tissues and differentially expressed between tumors and NATs(Gao et al., 2019; Ge et al., 2018).

**Figure 5.**
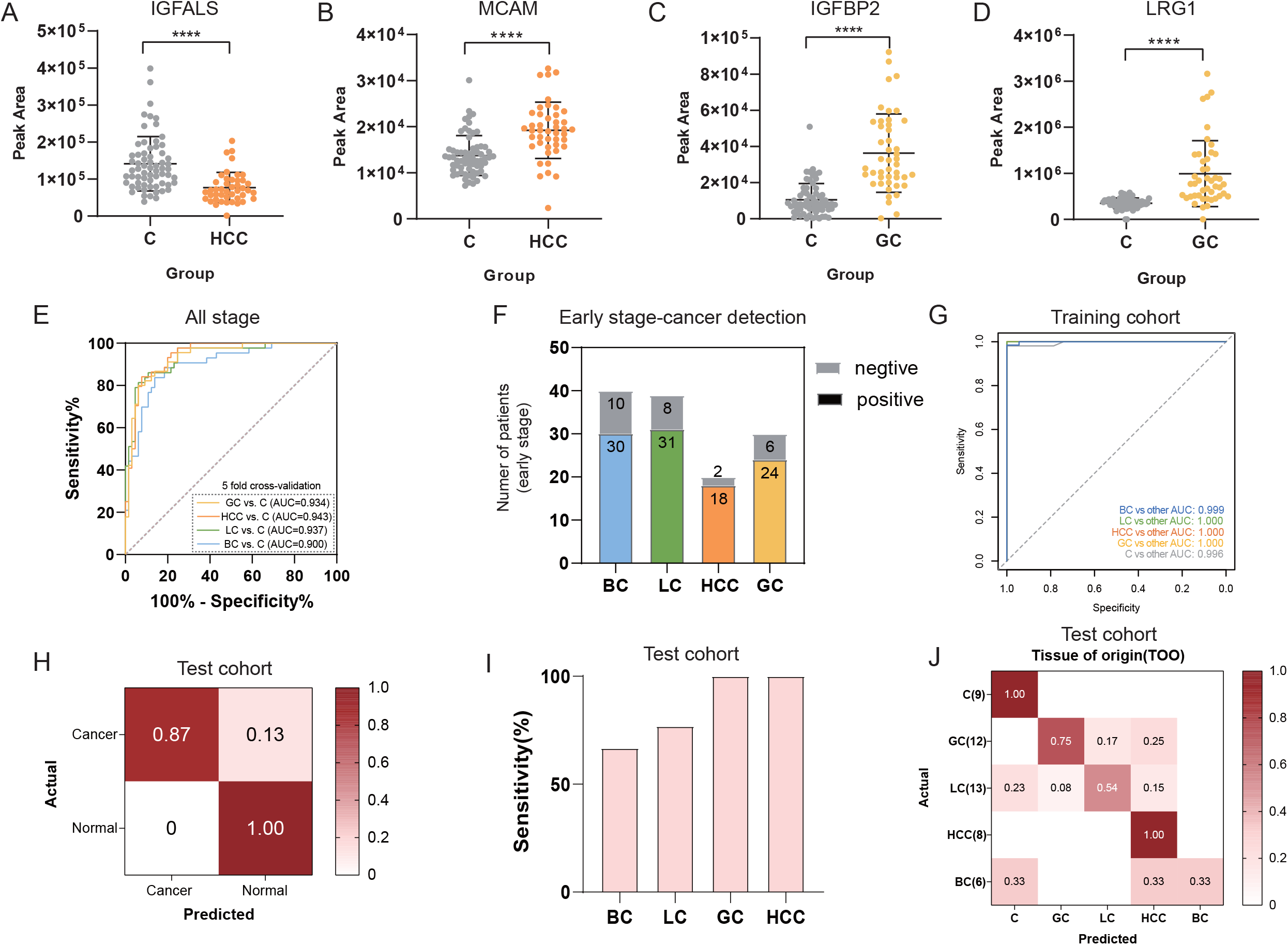
PPC-VDE assisted proteomics-based multi-cancer detection. (A-D) Scatter plots of the representative potential biomarkers. (E) ROC plots showing the performance of the diagnostic models in all-stage cancer detection of each cancer type. (F) Number of true positives in patients with early-stage cancer. (G) ROC plots of the five-binary classifier in the training cohort. (H) Confusion matrix showing the performance of multi-cancer detection model in the test cohort. (I) Sensitivity by cancer type in the test cohort. (J) Localization accuracy in the test cohort. *P-value was calculated via a two-tailed t-test. **** represents p-value < 0.0001.

Next, we constructed the diagnostic model based on logistic regression to classify the cancer patients and normal controls based on the pan-targeted proteomic data. The models can effectively distinguish cancers from normal controls, with the AUCs ranged from 0.900 (BC) to 0.943 (HCC) **(Figure 5E)**. The specificity ranged from 89.2% (BC) to 93.6% (LC) and the sensitivity ranged from 72.1% (BC) to 84.1% (HCC) **(Figure S5A-D)**. In particular, the sensitivity of the detection of early-stage cancer patients is not inferior to that of the late stages, ranging from 75% (BC) to 90% (HCC) (Stage 0-II, **Table S4, Figure 5F**). The results showed that the vast majority of early-stage cancer patients could be detected and thus the strategy is promising for early detection of cancer **(Figure 5F)**.

Finally, we tried to construct a multi-cancer detection model with the proteomic data and developed a customized probability-assisted multi-class random forest (PAR) model based on the training cohort of 240 samples **(Figure S5E)**. The rest 48 samples served as the independent test set. In the training cohort, we used 5-fold cross-validation, and the AUCs of five binary classifiers for HCC, GC, LC, BC, and C ranged from 0.996 to 1 **(Figure 5G)**. In the independent test cohort, the specificity of cancer detection was 100% and the overall sensitivity across cancer types was 87% **(Figure 5H)**. The sensitivity for each cancer type is shown in **Figure 5I**. GC and HCC had the highest sensitivity of 100%, followed by LC (76.9%) and BC (66.7%). The overall accuracy of tissue of origin (TOO) prediction was 73.8% in true positive cancer samples **(Figure 5J)**. In terms of each cancer type, the localization of HCC was the most accurate (100%), followed by GC and LC, while the accuracy of BC was only 50%. The low sensitivity and localization accuracy of BC may be attributed to the large proportion of early-stage (stage 0-II, 94%) patients involved in this study **(Figure S3A)**. Compared with the current DNA-based multi-cancer detection study **(Table S9)**(Cohen et al., 2018; Klein et al., 2021; Liu et al., 2020), our proteomics strategy showed a great sensitivity of cancer detection, with comparable localization accuracy **(Figure S5F, Table S9)**. This result preliminarily demonstrated the attractiveness of the proteomics-based multi-cancer detection model. In the following study, we will expand the sample size to further improve the diagnostic effectiveness.

## Discussion

Serum/plasma proteomics have been broadly applied to COVID-19(Messner et al., 2020; Shen et al., 2020), bacteremia(Wozniak et al., 2020), cancer(Dieters-Castator et al., 2019; Kim et al., 2021), cardiovascular diseases(Rueda et al., 2019) and so on(Sun et al., 2019; Xu et al., 2019). In these studies, untargeted DDA/DIA methods are typically used in the detection and discovery of differential proteins. Although they are non-biased, these untargeted approaches suffer from the overwhelming high-abundant proteins and their peptides, and unsatisfying repeatability and a requirement for complex data processing. In this study, we proposed the Cancer Serum Atlas supported pan-targeted proteomics strategy that can address the above problem and obtain the precise quantification of much cancer-secreted proteins in large cohort study. We firstly constructed Cancer Serum Atlas as a target protein/peptide bank (www.cancerserumatlas.com), over 2000 cancer-secreted proteins filtered from seven representative cancer types were included. These proteins have a high impact on primary tumors and can be directly used as target proteins in various serological studies. Then, we developed a SPA-PRM method featuring high accuracy, high sensitivity, high throughput, and wide linear range, enabling direct identification and quantification of hundreds of cancer-secreted proteins for large cohort studies, which accelerate the progress of biomarker discovery and clinical transformation. We noticed that Jarrod A. Marto group has just achieved comparable throughput with PRM-LIVE, an extensible, Python-based acquisition engine for the timsTOF Pro, which dynamically adjusts detection windows for reproducible target scheduling. (Zhu et al., 2021), It implies that our strategy is easy to implement in other laboratories with the support of the Cancer Serum Atlas. Researchers can quickly and efficiently find interested proteins and develop the SPA-PRM method with the information from the library, enables simultaneous biomarker discovery and validation. This library has already consisted of 2700 peptides corresponding to 2169 proteins, achieving deep coverage of cancer-secreted proteins **(Figure S2)**. Although these data were derived from seven cancer types, the library will be extended in future work and the Cancer Serum Atlas supported pan-targeted strategy can be applied to extensive cancer research.

We have proved that the Cancer Serum Atlas supported precise pan-targeted proteomics strategy is highly efficient for biomarker discovery and early detection of cancers. We further applied the strategy to construct a proteomics-based multi-cancer detection model. Multi-cancer detection would require high specificity, sensitivity, and accurate TOO identification. We demonstrated that four types of cancer can be detected through a protein-based blood test achieving high sensitivity (87.2%), specificity (100%), with 73.8% localization accuracy. Compared with the two promising multi-cancer detection approaches, CancerSEEK(Cohen et al., 2018) and Galleri(Klein et al., 2021; Liu et al., 2020), our approach has distinctive characteristics. The strategy needs an extremely low sample amount (240ng serum digest corresponding to 0.004 microliter serum), while the other two approaches need 10 milliliters of blood. The ability to use small samples amounts makes blood tests much less invasive and more clinically attractive. The diagnostic sensitivity of cancer (87.2%) is also far beyond the other two approaches (51.2% of Galleri and 62% of CancerSEEK). To our knowledge, this is the first time that multiple cancer could be simultaneously detected using MS-based proteomics technology. The localization accuracy is not as good as Galleri’s, probably due to the large proportion of the early-stage cancer patients (Stage 0-II, 87.5% of LC and 93.9% of BC, **Figure S3A**). We think that introducing more patients into the training cohort in the future may be helpful to find more disease-specific protein markers beneficial to the localization, even cancer detection. Taken together, the approach lays the conceptual and practical foundation for the proteomics-based blood test for multi-cancer detection. Expanding the sample size will be done in the following study to further improve the diagnostic effectiveness.

Although in the multi-cancer detection model, we only selected 4 types of cancer from which serum samples were initially collected, the representativeness of these 4 cancers is very strong. These 4 types of cancer have higher incidence and mortality worldwide **(Figure S1A)**. In the follow-up work, as more relevant studies are published, the database will be continually updated to introduce more cancer types, and we will also construct diagnostic models containing more cancer types in the follow-up research.

Collectively, our results provide a powerful tool for cancer research, and accelerates the progress of biomarker discovery, validation and clinical transformation. In addition, the proteomics-based multi-cancer detection model demonstrated the high sensitivity, specificity, and localization accuracy with extremely low sample consumption, which is clinically attractive.

## Materials and Methods

### Patients and clinical specimens

A total of 291 human serum samples were collected in Zhongshan Hospital (Shanghai) from Sep. to Nov., 2020. There are no biases in the selection of patients. The clinical characteristics including age, sex, cancer type, TNM stage, TNM grouping, histopathology and values of classical tumor markers were summarized in **Figure S3A-S3B** and **Table S4**. The medical conditions of controls is non-cancerous, apparently healthy people. This study was approved by the Research Ethics Committee from the Zhongshan hospital and Fudan University. Written informed consent was provided by all participants prior to participation. None of the patients underwent therapeutic measures such as radiation, chemotherapy, or surgery until sampling. All blood specimens were processed according to a standardized protocol, and serum aliquots were frozen until subsequent analysis.

### Chemicals and reagents

Mass spectra-grade trypsin was obtained from Promega (Promega, USA). Ammonium bicarbonate (ABC), dithiothreitol (DTT), Iodoacetamide (IAA), Acetonitrile (ACN, 99.9%), formic acid (FA,99.8%) and trifluoroacetic acid (TFA) were purchased from Sigma Aldrich (USA). The iRT kit was purchased from Biognosys (USA). All water used in the experiment was prepared using a Mill-Q system (Milipore, Bedford, MA).

### Selection of Targeted Proteins and Peptides

The differentially expressed proteins (DEP) were derived from the list of DEP provided in the corresponding literature **(Table S1)**. For studies that did not provide a DEP list, we performed DEP analysis (fold change>1.5 or fold change<0.67, p value < 0.05) using their raw data to obtain DEP lists. The primary tumor-derived DEP from literature mining were further filtered through bioinformatic prediction using SignalP-5.0 Server (http://www.cbs.dtu.dk/services/SignalP/) and Gene Ontology-cellular component (extracellular component, endoplasmic reticulum and Golgi apparatus) analysis (https://david.ncifcrf.gov/) **(Table S1-S2)**. For each protein, preferably two, at least one proteotypic peptide (PTPs) were selected from SRM Atlas (http://www.srmatlas.org/) or Skyline software based on the following criteria: (1) unique to a particular protein, (2) no peptide modifications, (3) fully tryptic. The proteins without eligible peptides were removed from the target protein list. An MRM transition list of peptides corresponding to targeted proteins was obtained from SRM Atlas(Kusebauch et al., 2016). Through preliminary screening in pooled serum using pseudo-targeted methods, the peptides with good signals were synthesized by Synpeptide Co.Ltd (Nanjing, China). Synthetic peptides were dissolved in 5% acetonitrile and stored at -80°C until use. Before MS analysis, all peptides were mixed, and the peptide mixtures were desalted by Sep-Pak C18 columns (Waters) and resuspended in 0.1% formic acid.

### MRM Assay and spectral library

The development and optimization of MRM assays for all standard peptides were assembled using a 6500 QTRAP mass spectrometer (SCIEX) using a 20-min liquid chromatograph (LC) gradient. For each peptide, the top ten transitions in the SRM Atlas were picked as preliminary MRM transitions. If more than one charge state was found, ten transitions of each charge state were selected and combined. MRM methods were generated by Skyline software. Collision energy (CE) optimization was set to use three steps on either side of the value predicted by the default equation, with the step size set to 3 V. In total, seven CE voltage values were considered for each fragment ion. The optimal CE is automatically selected by Skyline software according to the area of each transition chromatogram and the better responsive precursor with five or six most intense transitions for each peptide were manually picked out to generate MRM assay **(Table S3)** and spectral library as well as the next scheduled MRM experiment.

### 4D-MS/MS Assay and PRM spectral library

All standard peptides were mixed to generate the spectral library using timsTOF Pro mass spectrometer (Bruker) in the ddaPASEF mode using a 120-min LC gradient. The standard peptides’ spectral library for PRM was constructed using Mascot searched files from ddaPASEF analysis. For each peptide, compared with the spectral library, the matched peaks and the better responsive precursor were manually picked. Then, the method was exported to perform prmPASEF acquisition to generate the 4D-PRM spectral library.

### Batch design

After a careful check, 3 participants were excluded from the clinical cohort due to the unclear clinical characteristic or with a benign tumor. The validation experiment included 288 participants (**Table S4**) consisting of 52 Hepatocellular carcinoma (HCC), 57 Gastric cancer (GC), 56 Lung cancer (LC), 49 Breast cancer (BC), and 74 normal controls (C), which were randomly divided into a training set (n=240, **Figure 4A**) and an independent test set (n=48, **Figure 4A**). To minimize batch effects among the large cohort of serum, 240 samples of the training set were randomly distributed into 7 batches. Each batch contained 34 serum samples on average, one pooled serum sample as quality control (QC) for sample preparation, and one Hela digests as QC for the LC-MS system (**Figure 4A**). In the test set, 48 samples were analyzed following the above QC criteria. To ensure rigorous tests, the diagnoses were blinded during data acquisition and analyses.

### Protein digestion

The 1μL of each serum sample was diluted to 50μL with 50mM ABC. Proteins were reduced with 10mM DTT for 30min at 56°C, then alkylated with 20mM IAA for 30min in the dark at 37°C. Next, proteins were diluted with 70μL 50mM ABC at the estimated end concentration of 0.5 μg/μL. For digestion, the proteins were digested by trypsin at a mass ratio of 1:50 (enzyme: protein) for 12h at 37 °C on the shaker at 950rpm. Six microliters of 10% FA was added to quench the reaction. Digested peptides were cleaned-up with Sep PAK™ C18 column (Waters, USA), then centrifuged at 14000g for 30min and the supernatants were collected for future use.

### Targeted proteomics analysis in serum

The iRT standard was spiked into each sample before analysis. Standard peptides mixture was firstly analyzed using the optimized methods under full scan MRM mode with 120-min LC gradient to obtain the real-time retention time (RT). Then, the method was used to detect endogenous peptides in crude serum digests via scheduled MRM. For scheduled prmPASEF analysis, the standard peptides were also first analyzed using LC-MS/MS under the ddaPASEF scheme with a 60-min gradient to obtain the real-time RT and 1/K0 value. Then, the method was used to detect endogenous peptides in crude serum digests. In the process of the scheduled prmPASEF, the scanning time window of each peptide is set to 2 min, that is, 1 min before to 1 min after the pre-set retention time of the peptide. And the pre-set RT of a peptide is also real-time adjusted based on iRT standard during the experiment. Six transitions of each peptide were selected based on the rank in our spectral library for quantification.

### Quantitative performance evaluation

To evaluate the analytical performance of the developed prmPASEF assay, 10 standard peptides were added into an E. coli lysate and were separated using a 30-min separation gradient under prmPASEF mode. A dilution curve of nine concentration points, ranging from 0.25 to 100000 amole/μL was generated. Data from the dilution series were evaluated by linear regression analysis using triplicates for each dilution point. The Limit of detection (LOD) was defined as the lowest concentration at which the peak for the analyte is detected with an S/N ratio of 3. Limit of quantification (LOQ) was determined by serial dilution of the first calibrator until imprecision exceeded ±20%.

### LC-MRM Instrument parameters

The MRM analyses were carried out on a 6500 QTRAP hybrid triple quadrupole/linear ion trap mass spectrometer (SCIEX) interfaced with a UPLC system (Eksigent of SCIEX) using a 15-cm-long column (75-μm inner diameter; C18 3-μm silica beads) Mobile phases consisted of 0.1% (v/v) FA in water (phase A) and in acetonitrile (phase B) and the flow rate was set to 200nl/min. Gradient ranges and durations were as follows: 95% phase A over 1 min, 95-60% over 10 min, 60-20% over 0.2min, 20% over 0.8 min, 20-95% over 0.5 min and 95% over 7.5 min (20-min gradient); 92% phase A over 3 min, 92-75% over 67 min, 75-60% over 26 min, 60-20% over 4 min, 20% over 5 min, 20-92% 1 min and 92% over 14 min (120-min gradient). The spray voltage was 2300 V while the curtain gas was 25, gas 1 was 12 and the heat temperature of the ion source is 150 °C.

### LC-MS/MS and prmPASEF Instrument parameters

The LC-MS/MS and prmPASEF analysis were performed on a trapped ion mobility quadrupole time-of-flight (timsTOF Pro) mass spectrometer (Bruker) interfaced with a nano-HPLC (nanoElute, Bruker Daltonics) onto a 250mm × 75μm ID pulled emitter columns (IonOptiks) packed with 1.6μm C18-particles and heated at 50 °C in a column oven. Mobile phases consisted of 0.1% (v/v) FA in water (phase A) and in acetonitrile (phase B) and the flow rate was set to 200nl/min. Gradient ranges and durations were as follows: 2-22% phase B over 45 min, 22-37% over 5 min, 37-80% over 5 min, and 80% over 5 min (60-min gradient); 2-22% phase B over 90 min, 22-37% over 10 min, 37-80% over 10 min and 80% over 10 min (120-min gradient). For ddaPASEF acquisition, the range of mobility value was 0.6-1.6 Vs/cm^2^ (1/K0), the TIMS accumulation time and ion mobility separation time were both fixed at 50ms, and the covered m/z range was 300-1700 m/z. For prmPASEF acquisition in 288 serum, the TIMS accumulation time and ion mobility separation time were also fixed at 50ms. The range of mobility value was 0.65-1.35 Vs/cm^2^ (1/K0), and the covered m/z range was 300-1400 m/z. The ramp rate was 17.84 Hz and the collision energy varied from 20 to 59 eV according to the range of mobility (0.6-1.6 Vs/cm^2^). The capillary voltage was 1500 V while the dry gas was 3L/min, and the heat temperature of the ion source is 180 °C.

### Variable dimension expansion

The variable dimension expansion (VDE) algorithm was achieved with the Perl programming language written by our lab. The details have been described before(Xie et al., 2020).

### Machine learning analyses

#### Multi-cancer model training and evaluation on the discovery set

To accurately evaluate the performance of PPC-VDE data in multi-cancer detection and classification, we yield a matrix of 240 samples and 72900 features for feature selection. From 72900-DE features, we performed the orthogonal projections to latent structures-discriminant analysis (OPLS-DA) by SIMCA (Umetrics) to filter the features with top 1000 VIP scores in each one-versus-other classifier. The top 1000 features filtered by t-test were also used for feature selection. All the candidate features after the elimination of redundancy were input in the machine learning model for feature selection. The optimal number of features for each classifier was determined by the accuracy of cross-validation. Then, the selected features for each class versus other classes were used to train the five one-class-versus-other-classes binomial classifiers through 5-fold cross-validation.

#### Multi-cancer model evaluation on the independent test set

For each sample from the test set (48 samples), we estimated class probabilities for the C, GC, LC, HCC, and BC using one-versus-other classifiers trained before. The group with the highest score was determined as true.

### Luminex liquid suspension chip detection

Luminex liquid suspension chip detection was performed by Wayen Biotechnologies (Shanghai, China). The customized 4-plex kit containing AFP, CEA, NSE and OPN was used in accordance with the manufacturer’s instructions. In brief, serum samples (n=75) was incubated in 96-well plates embedded with microbeads for 1 h, and then incubated with detection antibody for 30 min. Subsequently, streptavidin-PE was added into each well for 10 min, and values were read using the Bio-Plex MAGPIX System (Bio-Rad).

### Statistical method and data analysis

The prmPASEF proteomic data were normalized by sum and missing values were imputed with the random number between 0 and the minimum value in each sample. A two-tailed t-test was used to identify the differentially expressed proteins. Skyline software (ver. 20.2, MacCoss Lab Software) was used to generate the methods and analyze the data from MRM, scheduled MRM, and prmPASEF experiments. Bioinformatics analyses were performed using SignalP 5.0 Server (http://www.cbs.dtu.dk/services/SignalP/), Gene Ontology Resource (http://geneontology.org/), Prism (ver.8.0.2, Graphpad Software Inc.), SIMCA (Umetrics) and STRING (ver.11.5, https://www.string-db.org/). Machine learning and ROC analysis were performed using R software. Correlation analyses were performed according to the previous study(Lapek et al., 2017).

## Data Availability

All data produced in the present work are contained in the manuscript.

## Compliance and ethics

The authors declare that they have no conflict of interest.

## Acknowledgments

This paper is dedicated to the memory of Professor Pengyuan Yang who passed away due to late-stage cancer on May 31, 2021.

The study was supported by the National Key Research and Development Program (2020YFE0202200 and 2017YFA0505001), the National Natural Science Foundation of China (81827901), and the innovative research team of the high-level local university in Shanghai, as well as the NHC Key Laboratory of Glycoconjugates Research.

## Author contributions

H.S., P.Y., A.H., and L.Z. planned and designed the project. Z.W., L.L., C.Y., and W.G. collected the serum samples and clinical characteristics. A.H., J.Z., and X.G. performed the sample preparation. L.Z., and A.H. performed mass spectrometry and data analyses. X.C. designed the website. H.S. and A.H. prepared the manuscript with the contributions from all authors.

