## Supplemental figures for "Cancer Serum Atlas supported precise pan-targeted proteomics enable multi-cancer detection"

‡This paper is dedicated to the memory of Professor Pengyuan Yang

### Table of Contents

### I.Supplemental figures and figure legends

Figure S1. Pan-cancer analysis of the proteomic data from literature and filtration of the cancer-secreted proteins.

Figure S2. Overview of the cancer-secreted proteins.

Figure S3. Data quality evaluation and quantitative statistics.

Figure S4. Protein-protein correlation (PPC) analysis and PPC based variable dimension expension (PPC-VDE).

Figure S5. Multi-cancer detection model development and performance comparison.

### II. Supplemental tables

Table S1. Literature mining

Table S2. Differentially expressed proteins (DEP) in primary tumor and predicted cancer-secreted proteins

Table S3. The optimized scheduled MRM methods for standard peptides

Table S4. Clinical characteristics of the cancer patients and controls

Table S5. Positive rate (sensitivity) by cancer type according to the clinical characteristics

Table S6. Quantification of 490 peptides corresponding to 400 cancer-related proteins using SPA-PRM method (after missing value estimation and normalization)

Table S7. Luminex liquid suspension chip analysis of tumor markers in 75 serum samples

Table S8. AUC comparsion analysis of classic tumor markers and pan-targeted proteomic data in Luninex cohort (N=75)

Table S9. Summary of 2 promising DNA-based multi-cancer detecion approach

I. **Supplemental figures and figure legends**

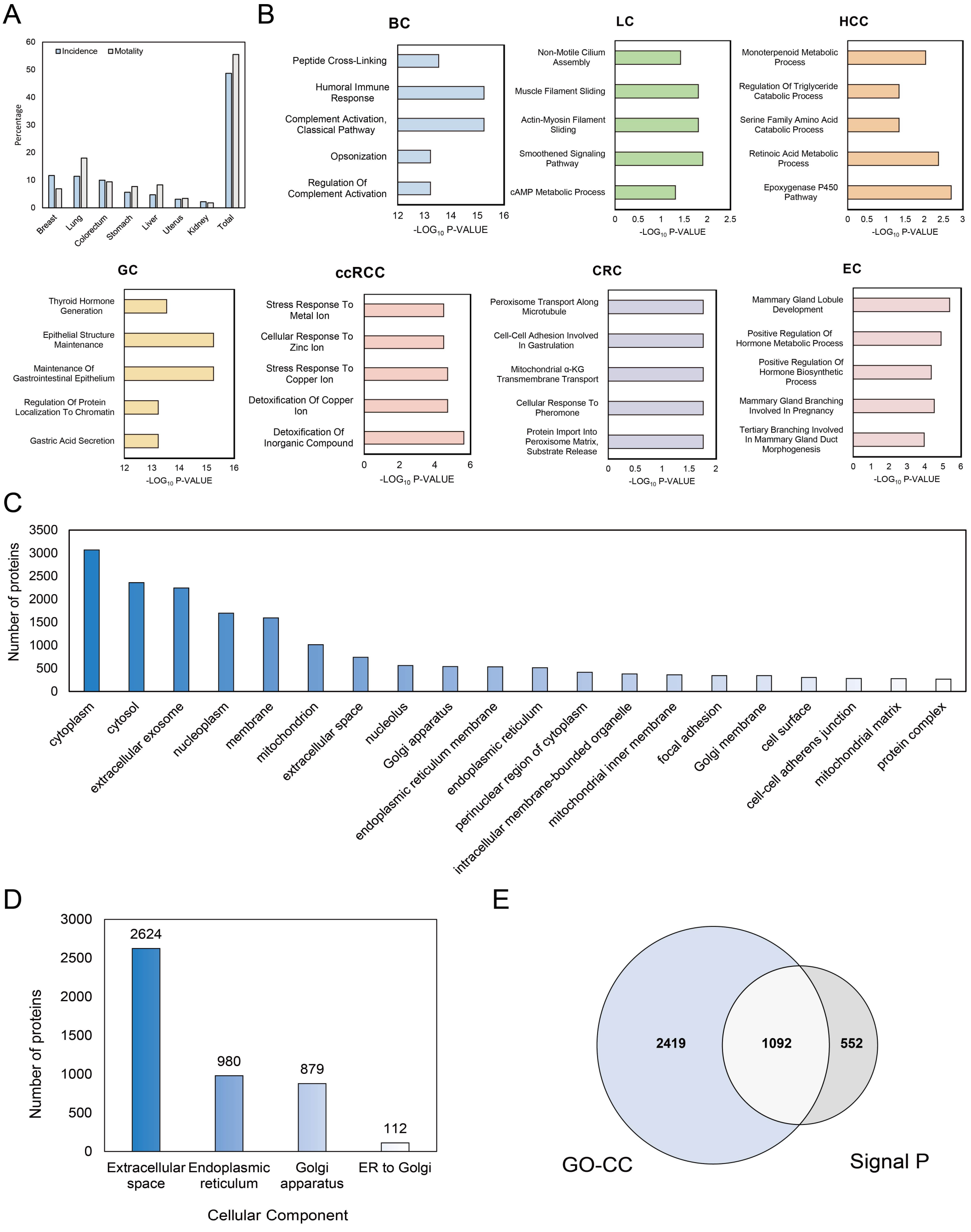


**Figure S1. Pan-cancer analysis of the proteomic data from literature and filtration of the cancer-secreted proteins.** (A) The incidence and morbidity of seven cancer types worldwide (http://gco.iarc.fr/). (B) Cancer-specific DEPs-related biological process. (C) Top 20 GO-CC terms of the 9096 DEPs. (D) Distribution of the cancer secretome analyzed by GO-CC. (E) Overlap of the cancer secretome analyzed by GO-CC and Signal P.

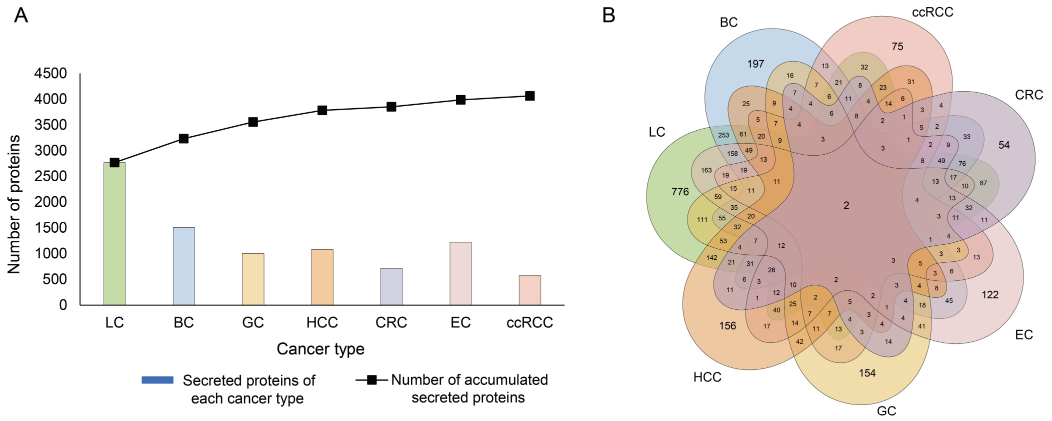


**Figure S2. Overview of the cancer-secreted proteins.** (A) Number of accumulated secreted proteins and the secreted proteins number of each cancer type. (B) Overlap of the cancer-secreted proteins of seven cancer types.

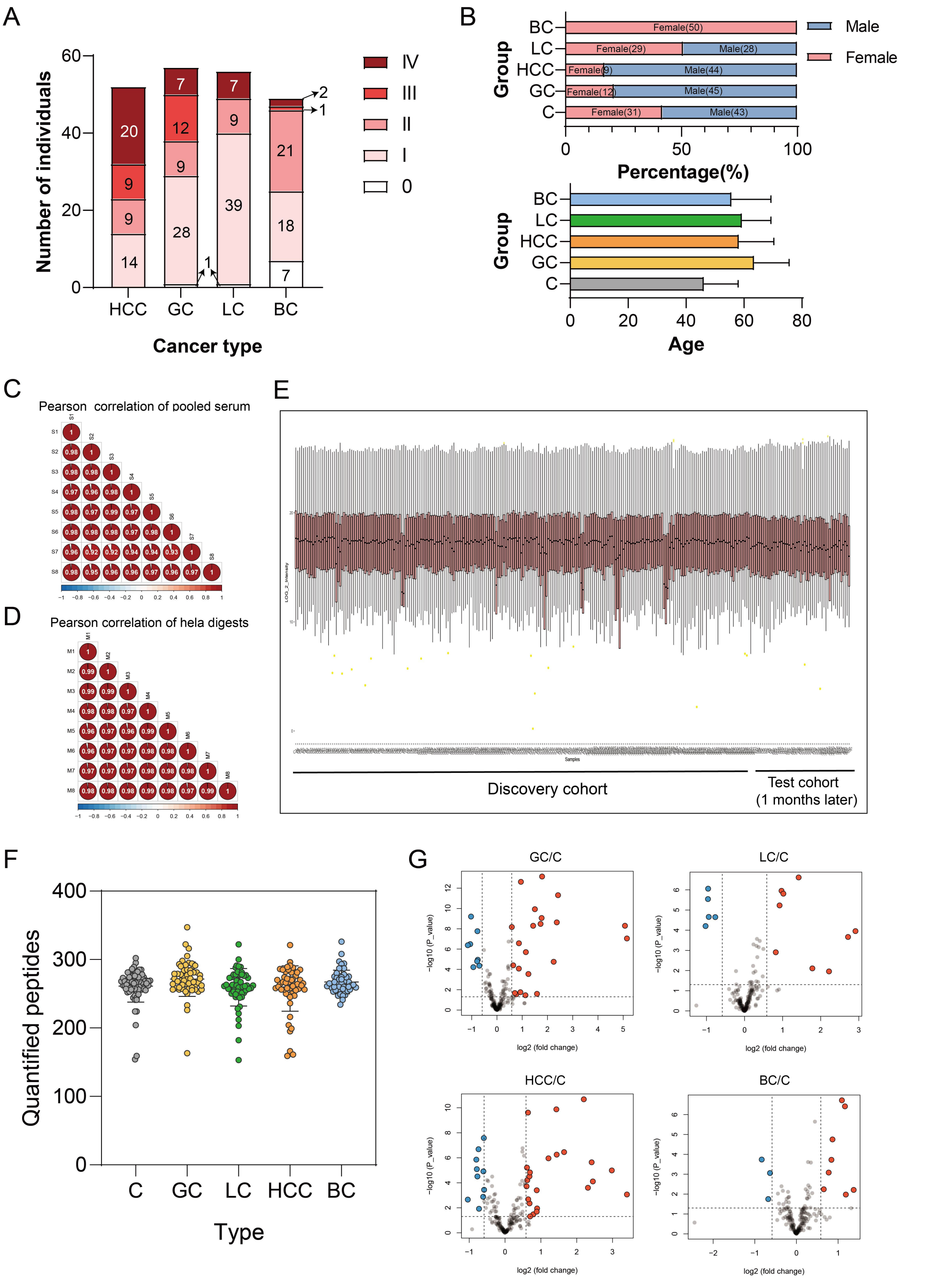


**Figure S3. Data quality evaluation and quantitative statistics**. (A-B) Summary of characteristics of the clinical cohort. Age distribution is calculated as mean with standard deviation. (C) Pearson correlation of eight pooled serum for quality control of sample preparation. (D) Pearson correlation of eight hela digests for quality control of mass spectrometry. (E) The total intensity distribution of 288 serum samples. (F) The number of peptides identified and quantified in each sample. (G) Volcano plots compare four cancer types of patient groups with control. Proteins with fold-change beyond 1.5 or below 0.67 with a p-value lower than 0.05 were indicated in the plot.

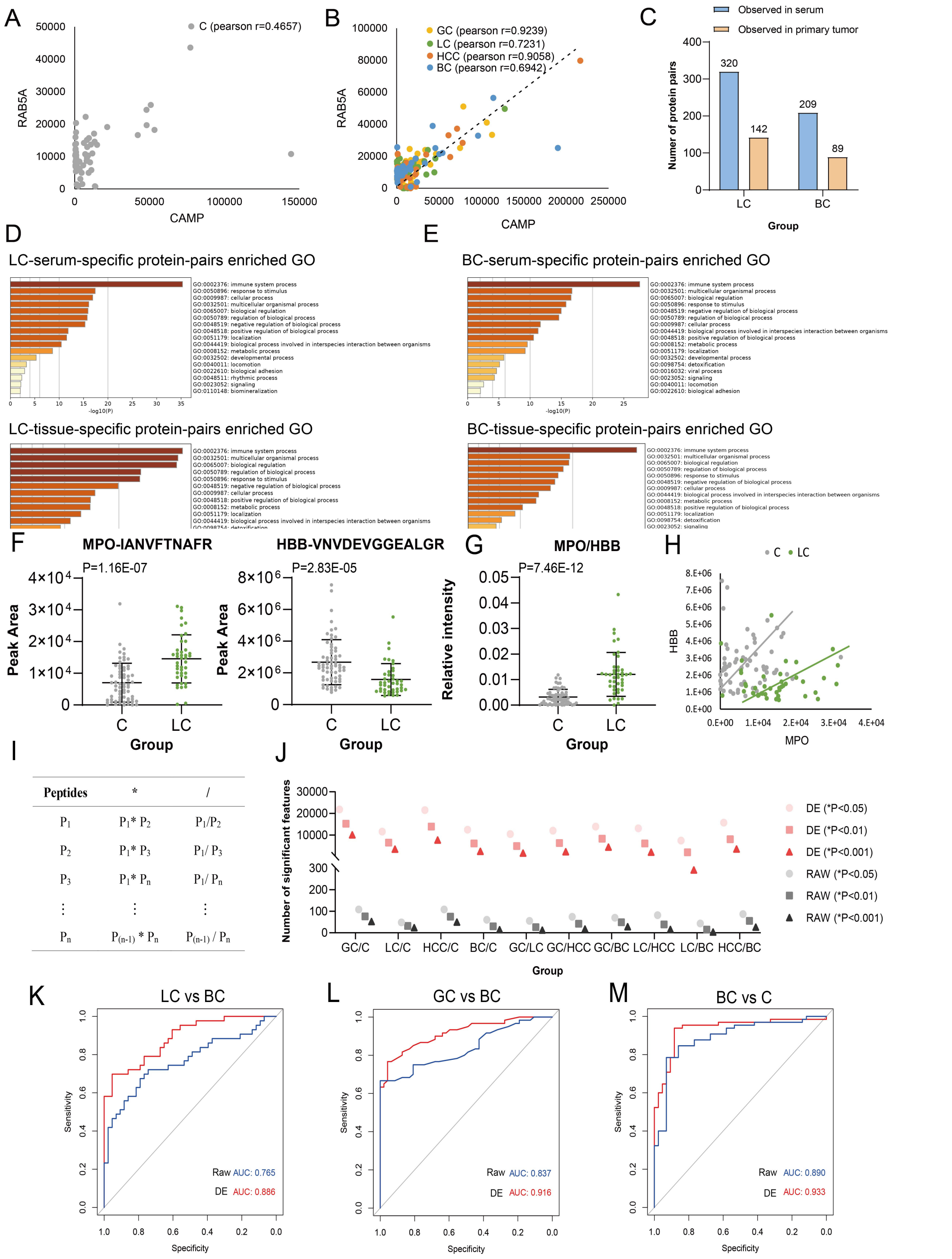


**Figure S4. Protein-protein correlation (PPC) analysis and PPC based variable dimension expension (PPC-VDE).** (A-B) The correlation of RAB5A and CAMP in normal controls and cancer groups. (C) The comparison of correlated protein-protein pairs observed in serum and primary tumor tissues (literature data). (D-E) The serum and tissue-specific protein pairs enriched GO-BP terms in LC and BC, respectively. (F-G) The peak area or relative intensity of unique peptide of MPO, HBB and MPO/HBB in C and LC groups. (H) The correlation of MPO and HBB in C and LC groups. (I) The general framework of PPC-VDE. (J) Comparison of significant features' between raw and DE data. (K-M) ROC plots showing the AUC value of raw and DE data.

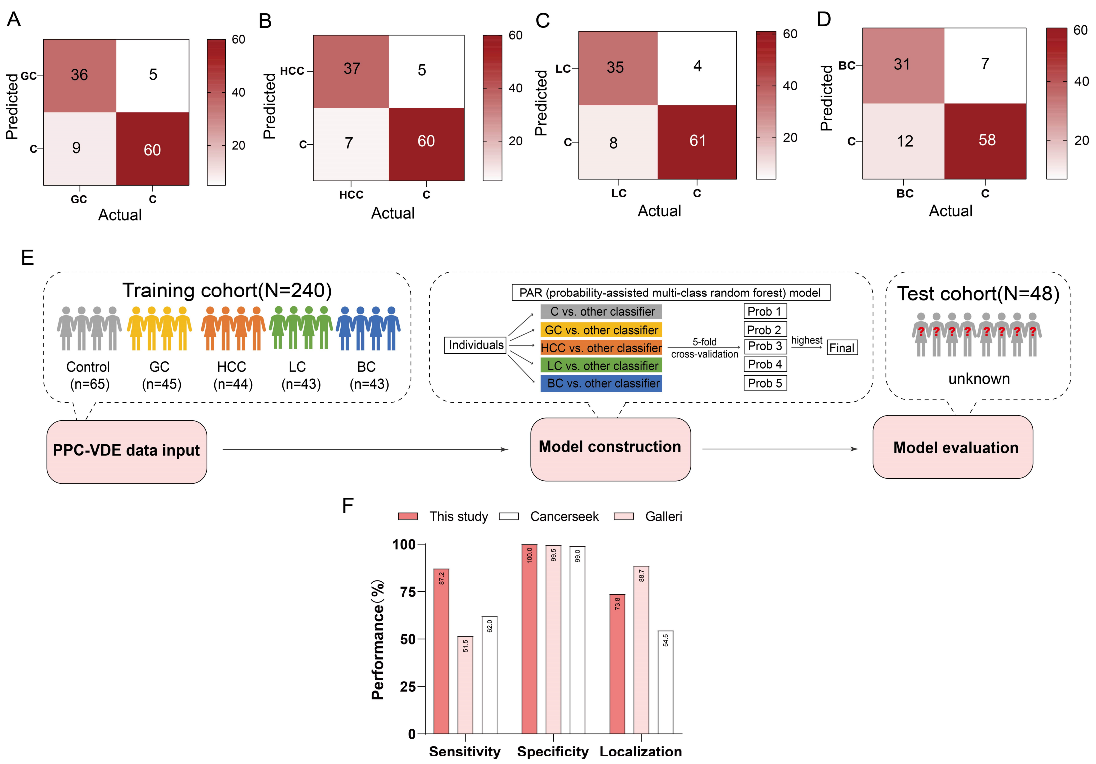


**Figure S5. Multi-cancer detection model development and performance comparison.** (A-D) Confusion matrix showing the performance of diagnostic models for detection of each cancer type. (E) Schematic of multi-cancer detection model construction and evaluation. (F) Comparison with current DNA-based multi-cancer detection study.
